# Hospitalist perspectives of available tests to monitor volume status in patients with heart failure: a qualitative study

**DOI:** 10.1101/2020.06.01.20119727

**Authors:** Anna M Maw, Carolina Ortiz-lopez, Megan A Morris, Christine Jones, Elaine Gee, Stephan Tchernodrinski, Henry Kramer, Benjamin T Galen, Amanda F Dempsey, Nilam Soni

## Abstract

Acute decompensated heart failure is the leading admitting diagnosis in patients 65 and older with more than 1 million hospitalizations per year in the US alone. Traditional tools to evaluate for and monitor volume status in patients with heart failure, including symptoms and physical exam findings, are known to have limited accuracy. In contrast, point of care lung ultrasound is a practical and evidenced-based tool for monitoring of volume status in patients with heart failure. However, few inpatient clinicians currently use this tool to monitor diuresis. We performed semi-structured interviews of 23 hospitalists practicing in 5 geographically diverse academic institutions in the US to better understand how hospitalists currently assess and monitor volume status in patients hospitalized with heart failure. We also explored their perceptions and attitudes toward adoption of lung ultrasound. Hospitalist participants reported poor reliability and confidence in the accuracy of traditional tools to monitor diuresis and expressed interest in learning or were already using lung ultrasound for this purpose. The time required for training and access to equipment that does not impede workflow were considered important barriers to its adoption by interviewees.

## Introduction

Acute decompensated heart failure is the leading admitting diagnosis in patients 65 and older with more than 1 million hospitalizations per year in the United States alone[1]. Current clinical practice guidelines recommend careful evaluation for signs of congestion and attainment of complete decongestion prior to discharge[1,2] because evidence of residual congestion at discharge is highly predictive of readmission[3,4]. However, it is well known that traditional tools to evaluate for volume overload including symptoms, physical exam findings and serum biomarkers (e.g. brain natriuretic peptide)[1,5] are of limited utility. This discrepancy between guideline recommendations and the limitation of traditional tests to evaluate and monitor volume status creates a conundrum for providers caring for patients hospitalized for heart failure.

Point-of-care lung ultrasound, which is performed and interpreted at the bedside by the treating clinician, offers a semi-quantitative and dynamic measure of pulmonary edema[6]. Recent randomized control trials have demonstrated the efficacy of lung ultrasound-guided diuresis in reducing urgent visits in patients recently hospitalized for heart failure[7] and length of stay in hospitalized patients[8]. Additionally, lung ultrasound is easy to learn and perform with excellent interrater reliability[9]. Despite robust data demonstrating its high accuracy[10], feasibility (easy to learn and perform)[11], patient centeredness (avoids radiation and improves patient satisfaction[12]), and endorsements by multiple society guidelines[13-15], few clinicians outside of emergency medicine and critical care use lung ultrasound routinely.

Given the gap between guideline recommendations for assessing volume status and the limited accuracy of traditional bedside tests to detect it, we sought to understand how hospitalists currently assess volume status, diuresis and discharge-readiness in patients hospitalized for heart failure as well as hospitalist perceptions and attitudes toward implementation of lung ultrasound as an evidenced-based tool in the assessment of patients with heart failure.

## Methods

### Study Sample and Setting

This qualitative study exploring the perspectives of hospitalists caring for patients with heart failure was deemed not Human Subjects research by the Institutional Review Board at University of Colorado. Eligibility criteria for enrollment included being a practicing physician who cares for hospitalized adults at an academic institution at least 1.5 months a year. Using snowball sampling we enrolled hospitalists from 5 academic medical centers located in geographically diverse regions around the United States. Recruitment was discontinued when ongoing preliminary analyses indicated thematic saturation: when no additional themes were emerging from the interviews.

### Data Collection

Interviews were conducted between June 2019 and March 2020. Interviews were conducted by phone or in person. The semi-structured interview guide was designed to explore hospitalists’ perceptions and attitudes toward current tools available to evaluate for volume status, parameters to determine discharge-readiness, utility of lung ultrasound and the factors that influence its adoption (see Supplementary Material). All interviews were conducted by a physician with prior experience with qualitative interviews[16] (AMM).

### Data Analysis

The interviews were audio recorded and transcribed verbatim. We used applied thematic analysis to guide our coding. Applied thematic analysis[17] is a systematic qualitative methodology well suited for capturing people’s experiences, views and perceptions of practical problems and is capable of providing an in-depth understanding of the tools used in the assessment of volume status and discharge readiness in patients hospitalized for heart failure as well the factors that inhibit or facilitate the use of lung ultrasound.

Both inductive and deductive coding was used to capture both expected and emerging themes. An initial deductive code list was developed from the interview guide which was then applied to the data. Additional inductive codes were identified through the review of the transcripts. Interview transcripts were coded and reconciled through consensus by least 2 research team members (CO, AMM). An outside auditor (MAM, a qualitative expert) oversaw the process to ensure credibility and confirmability of findings. Coded transcripts were analyzed within and across institutions to identify patterns of lung ultrasound implementation facilitators and challenges.

## Results

Interviews were conducted with 23 hospitalists at 5 academic institutions in diverse geographic locations. Interview length ranged from 35 to 60 minutes. Number of years in practice ranged from less than 1 to 18. Eleven participants were women. Ten interviewees reported no formal training in lung ultrasound. Nine reported brief training and 4 taught lung ultrasound to other faculty, trainees and students.

**Table 1.** Participant Demographics.

| Table 1. Participant Demographics. |  |
| --- | --- |
| Characteristics | n =23 (%) |
| Women | 11 (48) |
| Years in Practice | >1 to 18 |
| Experience with lung ultrasound |  |
| None | 10 (43) |
| Brief training | 9(39) |
| Instructor | 4(17) |
| Geographic Area |  |
| Midwest | 1 (4) |
| Northeast | 9 (39) |
| Rocky Mountain | 11(48) |
| South | 1 (4) |

### Themes

Hospitalists reported the poor reliability of traditional tools to assess changes in volume status such as physical exam techniques, serial weights, and a recordings of liquids administered and urine volume (I/Os). In contrast, most interviewees felt lung ultrasound would be useful in the assessment of volume status. All expressed interest in learning to use or were already using lung ultrasound to monitor diuresis in their patients hospitalized for heart failure. Those not yet trained reported the time required to learn to perform lung ultrasound as one of the major barriers to its adoption. Interviewees also noted the time required to retrieve the ultrasound prior to use as an important barrier to its regular application. There were no differences in themes across sites or hospitalist skill at lung ultrasound (Table 2).

**Table 2.** Themes.

| Table 2. Themes. |  |  |
| --- | --- | --- |
| Themes | Description | Exemplar Quotes |
| **Reliability of Traditional Tools** | Traditional tools are unreliable | *“The challenge is weights in the hospital fluctuate in a manner that make me suspect measurement inaccuracy rather than change in patient status.”* |
| **Determining Euvolemia** | Difficult to determine with traditional tools | *“He looked dry to us, and to the cardiologist, as well, on exam. But his wedge was 24.”* |
| **Discharge readiness with regard to volume status** | Euvolemia is not necessarily a goal of hospitalization | *“But I don't actually think euvolemia is necessary in every patient.”* |
| **Utility of lung ultrasound** | Perceived to be helpful in determination of volume status | *“Lung ultrasound really has the promise of safe, accurate, radiation-free, fast diagnostics.”* |
| **Desire to perform lung ultrasound** | Interviewees not already trained expressed a desire for training | *“I would love to be integrating that into my practice. I wish that I had that training.”* |
| **Factors that influence adoption of lung ultrasound** | Time required to access equipment | *“There's a huge effect of convenience factor because when I'm on med two and I have an ultrasound around my shoulder, I use it a heck of a lot more than on any other service.”* |
|  | Adequate training | *“When you think about ultrasound I think of user reliability. That's the biggest thing I worry about.”* |
|  | Billing | *“if we could bill for it, maybe they could roll that right back into our development in this area.”* |
|  | Institutional Culture | *“There's a certain barrier with new adoption of tools. I think that requires a little bit of a cultural shift”* |

### Reliability of current tools

Many hospitalists noted the limitations of the accuracy of available tools to evaluate volume status and monitor diuresis. Hospitalists described that the different sources of data about volume status frequently conflict with each other, leading to uncertainty:

### Participant 1

“That can definitely be a challenge. I think we all order the strict I’s and O’s, the daily weights, things like that. But then I see the weight sometimes and they’re net negative two liters from the night before, but their weight is up and I don’t know which one is right.”

### Determining Euvolemia

Interviewees expressed difficulty in determining euvolemia with the traditional tools of physical exam, weight, recordings of I/Os.

#### Participant 2

“I think I would look at urine output. If I feel like the weights have been somewhat reliable, I’ll look at the weights. I’ll ask the patient how they feel. I’ll ask them to look at their legs and see if they feel like they’re back to normal or if they’re too big. A lot of times I’ll rely on the patient to tell me what’s normal for them. I don’t follow serial BNPs.”

#### Participant 3

“I think one of the other challenges about CHF is that I think it’s easy to fall into this trap where, ‘Oh, I see one sign where they’ve hit the goal and now I’m done’, when really the truth about euvolemia is you need to triangulate it based on multiple data points. I don’t see that being done a lot.”

### Discharge readiness with regard to volume status

Many interviewees felt that euvolemia was not necessarily the goal of hospitalization and that some patients could be discharged to complete diuresis as an outpatient if they had adequate follow-up and social support.

#### Participant 6

“I find one of my challenges with heart failure admissions is trying to figure out the tension between the need to keep length of stay as short as possible and an awareness that patients who are no longer floridly decompensated are generally not back to true dry weight, true homeostasis for a period of a number of days. One of the more challenging aspects of clinical decision making in heart failure is what to do with that window between no longer being acutely symptomatic, decompensated heart failure, and once they are truly back to a maintenance diuretic regimen at dry weight.”

#### Participant 10

“I think some patients that can achieve good diuresis with oral regimen that I know is going to work, I know they have good follow-up and a good understanding of how to push that. I think that some patients can successfully do that at home.”

### Utility of lung ultrasound

Interviewees indicated they felt lung ultrasound was useful for improving the timeliness of diagnosis and appropriate treatment in addition to conserving resources.

#### Participant 15

“I think that the real benefit in the acute hospitalized patient who has acute onset hypoxemic failure or acute onset of dyspnea is really where it’s a huge game changer because these are cases where you’re wondering, ‘Did they aspirate? Do they have a PE? Or is this flash pulmonary edema?….But I think that the flash setting, when you have a case of someone who’s acutely dyspneic, and you evaluate them with ultrasound, it really spares them a lot of delay in the right care and it also spares a lot of unnecessary treatment.”

### Desire to use lung ultrasound

Interviewees indicated a strong desire to learn to perform lung ultrasound but acknowledged barriers to this including time required to locate the ultrasound, adequate training and institutional culture.

#### Participant 2

“I’d like to think that all eager and enthusiastic physicians won’t find learning that new modality a barrier. I don’t get that sense…. People want to learn it. I would be willing to learn it.”

### Factors that influence the adoption of lung ultrasound

#### Time required to use the ultrasound

The time required to use the ultrasound because of barriers to access was a frequently mentioned barrier by physicians. Although most participants reported their practice setting had ultrasound machines available, they usually required more time to access than was practical and for that reason was less often used.

#### Participant 19

“I think the only factor that prevents most people from using it is having their own personal ultrasound device….If there’s only two or three ultrasounds, even if there’s a community ultrasound in the hospitalist work room, no one’s going to take the time, if it’s a shared device, to go use it on all their patients.”

### Adequate training

Interviewees expressed concern that lack of adequate training may limit effective use of lung ultrasound.

#### Participant 13

“The biggest one is how much confidence they have in their skill, I would say. If they don’t feel like they can trust their either ability to acquire images or interpret them, then ultrasound won’t be very helpful.”

#### Participant 1

“The person who uses the ultrasound every day versus the person who does it every once in a while they may not be interpreting what they’re seeing correctly. Which is something I worry about. Versus somebody who is good at using it and can be like, ‘Oh yes’ and teaches other people, this is what I’m looking for.”

### Billing

The ability to bill was considered a facilitator to lung ultrasound implementation.

#### Participant 2

“Yeah, I think if that (billing) were in place, then the division would have a lot more interest in getting us trained and supporting our use of it and maybe even giving us the tech that we would need to make it easy

### Institutional culture

#### Participant 20

“The whole organization needs to recognize lung ultrasound as much as they would a lab finding or an imaging finding that they can all look at it and interpret themselves.”

## Discussion

To our knowledge, this is the first qualitative study to capture hospitalist practices/perceptions, and attitudes relating to the assessment, monitoring and management of volume status in patients hospitalized for heart failure. Our findings suggest hospitalists perceive traditional tests used to assess volume status and monitor diuresis as difficult to use and unreliable. They also suggest that some hospitalists do not consider euvolemia a goal of hospitalization and that given enough social support and follow-up, some patients can safely continue to diuresis as an outpatient. This management style is in contrast to current professional guidelines and should be further studied.

Our findings also demonstrate that study participants perceived lung ultrasound as a clinically useful tool and desired further training to learn how to use it to guide heart failure management. Interviewees felt factors influencing adoption of lung ultrasound included the time required to access the equipment, time to train, ability to bill and institutional culture. Although most providers reported their clinical environments had at least one ultrasound machine, the act of having to retrieve the ultrasound for every use was a significant enough disruption in workflow to be an important barrier to its use. Many participants pointed out that given the rapidly improving image quality of hand-held ultrasounds in conjunction with their decreasing price, access to ultrasound machines will likely pose less of a barrier in the years to come as the majority of providers may acquire their own personal devices, they carry with them. Many interviewees also reported finding the time to train was a barrier to using lung ultrasound. This suggests lung ultrasound educators should consider creating curricula that better meet the needs of busy clinicians. This may require a nontraditional course structure that offers more flexibility in scheduling such as online didactics and supervised scanning of patients while the learner is on service.

Interviewees also noted systems level factors that may influence adoption including billing and institutional culture. We plan to further explore system level factors with interviews of stakeholders in roles at multiple levels within a healthcare system including hospital administrators, radiologists, information technologists and payers.

### Limitations

There are several limitations to our study. The participants in this study all practiced in academic settings and many had already integrated lung ultrasound into their care of patients hospitalized for heart failure. This may limit the generalizability of our results as the majority of hospitalists work in community settings and may not have received training in lung ultrasound.

With regard to factors that may influence implementation of lung ultrasound, this study would have been improved by gaining the perspective of stakeholders at multiple levels of the institution in the assessment of factors that influence adoption at a systems level. We plan to capture these perspectives in future work.

## Conclusions

Our findings suggest academic hospitalists recognize the limitations of current tools to accurately assess residual congestion and are interested in learning how to perform lung ultrasound to improve their bedside assessment of volume status. Remaining barriers to adoption of lung ultrasound include time for training and access to equipment that won’t impede workflow.

## Data Availability

All data referred to in the manuscript is available

## Author Contributions

Conceptualization, AMM. and MAM.; Methodology, CDJ and MAM.; Formal Analysis, AMM, and MAM.; Investigation, AMM, CO, EG, ST,HK and BTG.; Data Curation, AMM, MAM, CDJ, AFD and NJS.; Writing—Original Draft Presentation, AMM; Writing - Review & Editing AMM, CO, MAM, CDJ, EG,ST,HK, BTG,AFD, and NJS.; Supervision, MAM, AFD, and NJS.

